# Comparison of antibody immune responses between BNT162b2 and mRNA-1273 SARS-CoV-2 vaccines in naïve and previously infected individuals

**DOI:** 10.1101/2021.10.05.21264550

**Authors:** Duaa W. Al-Sadeq, Farah M. Shurrab, Ahmed Ismail, Fathima Humaira Amanullah, Swapna Thomas, Nader Aldewik, Hadi M. Yassine, Hanan F. Abdul Rahim, Laith Abu-Raddad, Gheyath K. Nasrallah

**Affiliations:** Biomedical Research Center, Qatar University, Doha 2713, Qatar; College of Medicine, QU Health, Qatar University, P.O. Box 2713, Doha, Qatar; Laboratory Section, Medical Commission Department, Ministry of Public Health, Doha, Qatar; Department of Biological and Environmental Sciences, College of Arts and Science, Qatar University, P.O. Box 2713, Doha, Qatar; Clinical and Metabolic Genetics Section, Pediatrics Department, Hamad General Hospital (HGH), Women’s Wellness and Research Center (WWRC), Interim Translational Research Institute (iTRI), Hamad Medical Corporation (HMC), College of Health and Life Science (CHLS), Hamad Bin Khalifa University (HBKU), Doha, Qatar, P.O. Box 3050, Doha, Qatar; Department of Biomedical Science, College of Health Sciences, QU Health, Qatar University, P.O. Box 2713, Doha, Qatar; College of Health Sciences, QU Health, Qatar University, P.O. Box 2713, Doha, Qatar; Infectious Disease Epidemiology Group, Weill Cornell Medicine□Qatar, Cornell University, Qatar Foundation – Education City, Doha, Qatar; World Health Organization Collaborating Centre for Disease Epidemiology Analytics on HIV/AIDS, Sexually Transmitted Infections, and Viral Hepatitis, Weill Cornell Medicine–Qatar, Cornell University, Qatar Foundation – Education City, Doha, Qatar; Department of Healthcare Policy and Research, Weill Cornell Medicine, Cornell University, New York, United States of America

**Keywords:** Pfizer-BNT162b2, Moderna-mRNA-1273, S-RBD IgA, S-RBD IgG, Immune response

## Abstract

Two mRNA vaccines, Pfizer-BNT162b2 and Moderna-mRNA-1273, were granted the US Food and Drug Administration Emergency Use Authorization for preventing COVID-19. However, little is known about the difference in antibody responses induced by the two mRNA vaccines in naïve and individuals with a previous history of infections (PI group). Therefore, we investigated the levels of anti-S-RBD total antibodies (IgM, IgA, and IgG), anti-S-RBD IgG, and anti-S-RBD IgA in these two groups 1-13 (median=6) weeks following administration of two doses of mRNA-1273 or BNT162b2 vaccines. Results showed that in naïve-vaccinated group, the mRNA-1327 vaccine induces significantly higher levels of S-RBD total antibodies (3.5-fold; p<0.001), S-RBD IgG (2-fold-p<0.01), and S-IgA (2.1-fold, p<0.001) than the BNT162b2 vaccine. In the PI-vaccinated group, both vaccines produce significantly higher S-RBD total antibodies level than those of the naïve-vaccinated group. The PI group produced a higher level of S-RBD IgG than the naïve-BNT162b2 (p=0.05) but not more than the naïve-mRNA-1273 (p=0.9) group. Interestingly, the PI-vaccinated group produced a comparable level of IgA ratio to the naïve-mRNA-1273 group but significantly higher than the naïve-BNT162b2 group (1.6-fold, p<0.001). Our results showed that the mRNA-1327 vaccine is more immunogenic and induces a greater antibody response than the BNT162b2 vaccine.

## 1. Introduction

Since the beginning of the COVID-19 pandemic, over 226 million people have been infected with the Severe Acute Respiratory Syndrome Coronavirus 2 (SARS-CoV-2), with more than four million deaths attributed to COVID-19 [1]. To combat the wide spread of SARS-CoV-2, major vaccination campaigns have been launched worldwide, with 3.9 billion vaccine doses provided to date [2]. In December 2020, the US Federal Food and Drug Administration (FDA) approved emergency authorization use (EAU) to Pfizer-BNT162b2 and Moderna-mRNA-1273COVID-19 vaccines.

Recent clinical studies and controlled trials have demonstrated the efficacy of the FDA-approved COVID-19 vaccines. The developed mRNA vaccines BNT162b2 (Pfizer-BioNTech) and mRNA-1273 (Moderna) were previously reported as safe vaccines with an efficacy rate of more than 94% [3, 4]. Recently we have demonstrated that the effectiveness of the mRNA-1273 shows similar levels and patterns of protection to the BNT162b2 vaccine. Still, the mRNA-1273 appears to be more robust against the SARS-CoV-2 B.1.351 variant and provides greater protection than the BNT162b2 [5]. However, the antibody immune response to different mRNA COVID-19 vaccines has not been extensively studied, emphasizing the need to evaluate the FDA-authorized vaccines’ durability and comparative efficacy. Therefore, this study aims to compare the antibody immune responses between the mRNA-1273 and BNT162b2 SARS-CoV-2 vaccines after administering two doses in naïve and previously infected participants. Qatar is currently administering the BNT162b2 and mRNA-1273 vaccines. Both vaccines have been approved for emergency use by the Department of Pharmacy and Pharmaceutical Control in the Ministry of Public Health.

## 2. Material and Methods

### 2.1 Ethical approval and sample collection

Randomized participants who received two doses of either BNT162b2 or mRNA-1273 vaccine were eligible for inclusion. A total of 289 samples were collected from staff and students of Qatar University, the national university Qatar, with different age groups and nationalities. Peripheral blood was collected 1-13 (BNT162b2 median=6, mRNA-1273 median=5, PI median=6) weeks following the administration of the second dose of vaccine. Participants were either naïve or previously infected (PI) with SARS-CoV-2. The study was reviewed and approved by the Institutional Review Board at Qatar University (QU-IRB 1537-FBA/21). Plasma was separated from venous whole blood collection and stored at -80 °C until performing the immunoassays testing. Demographic information was collected through a self-administered questionnaire. In addition, the questionnaire included questions regarding the previous history of infection.

### 2.2 Serology testing

Serological testing was done using the automated analyzer CL-900i® from Mindray Bio-Medical Electronics [6-8] using two chemiluminescence immunoassays to detect the vaccine-induced antibodies: (i) The SARS-CoV-2 S-RBD IgG (catalog No. SARS-CoV-2 S-RBD IgG122, Mindray, China), the cut off index for the kit is ≥10-1000 BAU/mL, and (ii) The anti-S-RBD SARS-CoV-2 total antibodies (IgG, IgA, and IgM) (Catalog No. SARS-CoV-2 Total Antibodies 122, Mindray, China) with positive cut off index of ≥10-2000 AU/mL. All samples with readings higher than the reference range were diluted with phosphate-buffered saline (PBS). In addition, the Euroimmun anti-SARS-CoV-2 IgA ELISA (Catalog No. EI 2606-9601 A, Germany) was used to measure the anti-S1 antibody levels [9]. The IgA ratio was calculated by dividing the extinction of the sample by the calibrator. Ratios ≥ 1.1 are considered positive, <0.8 are negative, and ≥0.8 to <1 are borderline. Some samples were not tested for IgA (40/289) because of limited plasma volume. All tests were carried out according to the manufacturers’ instructions.

In addition to the information collected by the questionnaire about the previous history of infection, we used nucleoprotein-specific IgG (anti-N IgG) to denote prior SARS-CoV-2 exposure. That is, the naïve, but not the previously infected volunteers, do not produce IgG to the N protein. Thus, the previous infection is defined as anti-N positivity and/or reported history of positive polymerase chain reaction results on the nasopharyngeal swab. All samples were tested for the presence of anti-N SARS-CoV-2 IgG using the Architect automated chemiluminescent assay (Abbott Laboratories, USA) according to the manufacturers’ instructions [8].

### 2.3 Statistical analysis

Data were analyzed using GraphPad Prism 9.2.0. (San Diego, CA, USA). Results in the graphs are plotted as mean values with standard deviation (SD). One-way ANOVA tests were performed to compare the groups, and p values <0.05 were considered statistically significant. In all graphs, *p ≤ 0.05, **p ≤ 0.01, and ***p ≤ 0.001.

## 3. Results

### 3.1 Participant characteristic

Participants’ characteristics are described in Table 1. A total of 289 mRNA naïve vaccinated and PI vaccinated volunteers participated in this study. Of those, 218 were BNT162b2 naïve vaccinated, 45 naïve mRNA-1273 vaccinated, and 26 PI vaccinated participants (23 with BNT162b2 vaccine and 3 with mRNA-1273 and). Further details about participants are provided in Table S1 and S2.

**Table 1.** Demographic characteristics of the study sample (*n* = 289).

| Characteristic |  | BNT162b2 (%) |  | mRNA-1273 (%) |  |
| --- | --- | --- | --- | --- | --- |
|  |  | Naïve | PI | Naïve | PI |
| Gender | Male | 114 (52.29) | 14 (60.86) | 23 (51.11) | 1 (33.33) |
|  | Female | 104 (47.70) | 9 (39.13) | 22 (48.88) | 2 (66.66) |
|  | Total | 218 | 23 | 45 | 3 |
| Age<br>(years) | <30 | 71 (43.29) | 8 (34.78) | 26 (57.77) | 1 (33.33) |
|  | 30–50 | 104 (50.24) | 14 (60.86) | 16 (35.55) | 2 (66.66) |
|  | >50 | 32 (15.45) | 1 (4.34) | 2 (4.44) | - |
|  | Unknown | - | - | 1 (2.22) | - |
|  | Total | 218 | 23 | 45 | 3 |

### 3.2 Antibody immune response assessment following vaccination

#### 3.2.1 S-RBD total antibodies response

All naïve-BNT162b2, naïve-mRNA-1273, and PI vaccinated group has positive total antibodies response (Figure 1A), with mean levels of 4.6×10^3^ AU/mL (95%CI: 3.0-6.2×10^3^), 1.6×10^4^ AU/mL (95%CI: 6.1×10^3^-2.7×10^4^), and 3.0×10^4^ AU/mL (95%CI: 1.4-4.6×10^4^), respectively. The naïve-mRNA-1273 vaccinated group produced significantly higher total antibodies level than the naïve-BNT162b2 (4-fold, p<0.01). The PI vaccinated group produced significantly higher levels of total S-RBD total antibodies compared to the naïve-BNT162b2 (6.5-fold; p<0.001) and naïve-mRNA-1273 (1.9 fold, p<0.05) vaccinated group.

**Figure 1.**
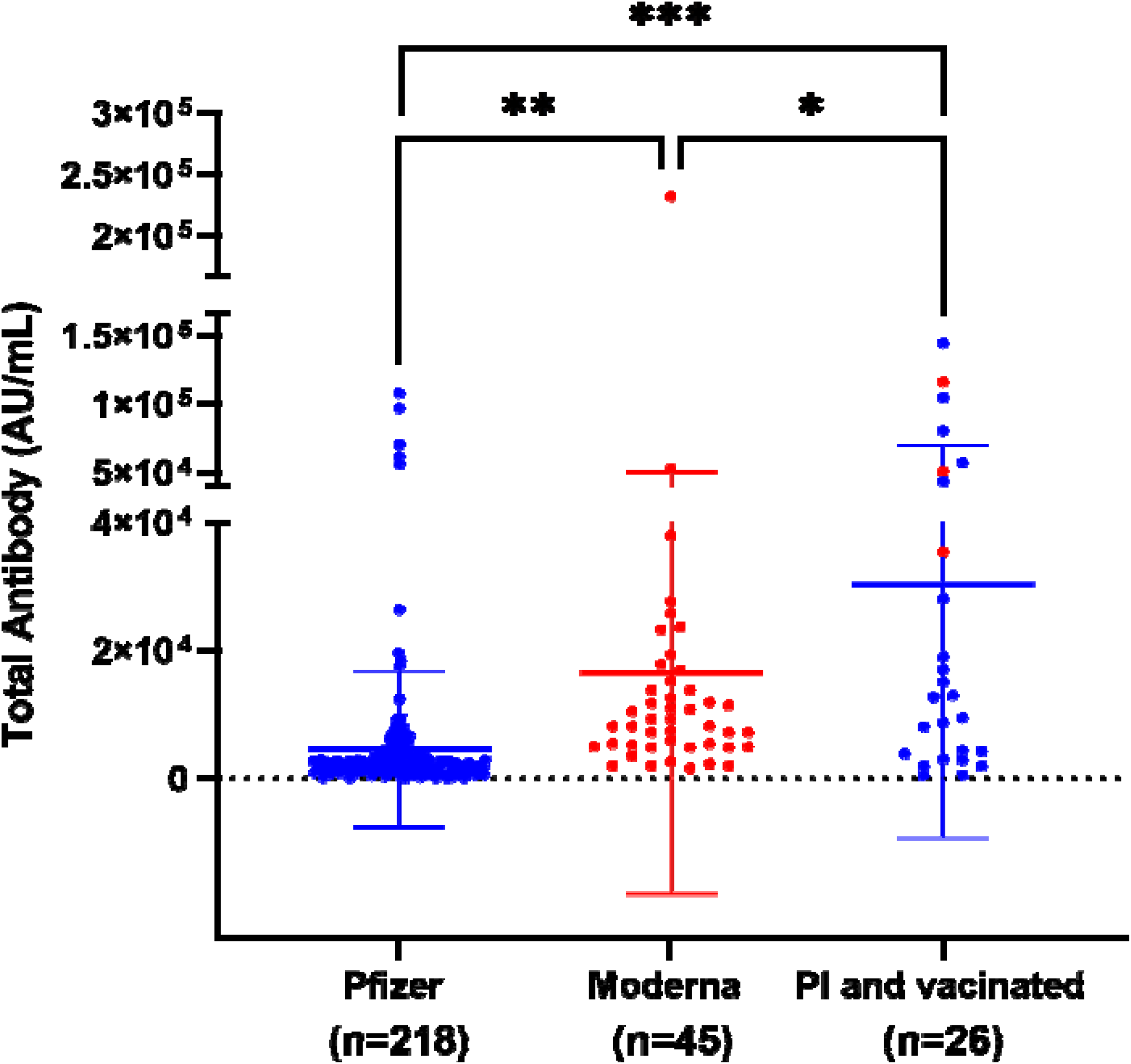
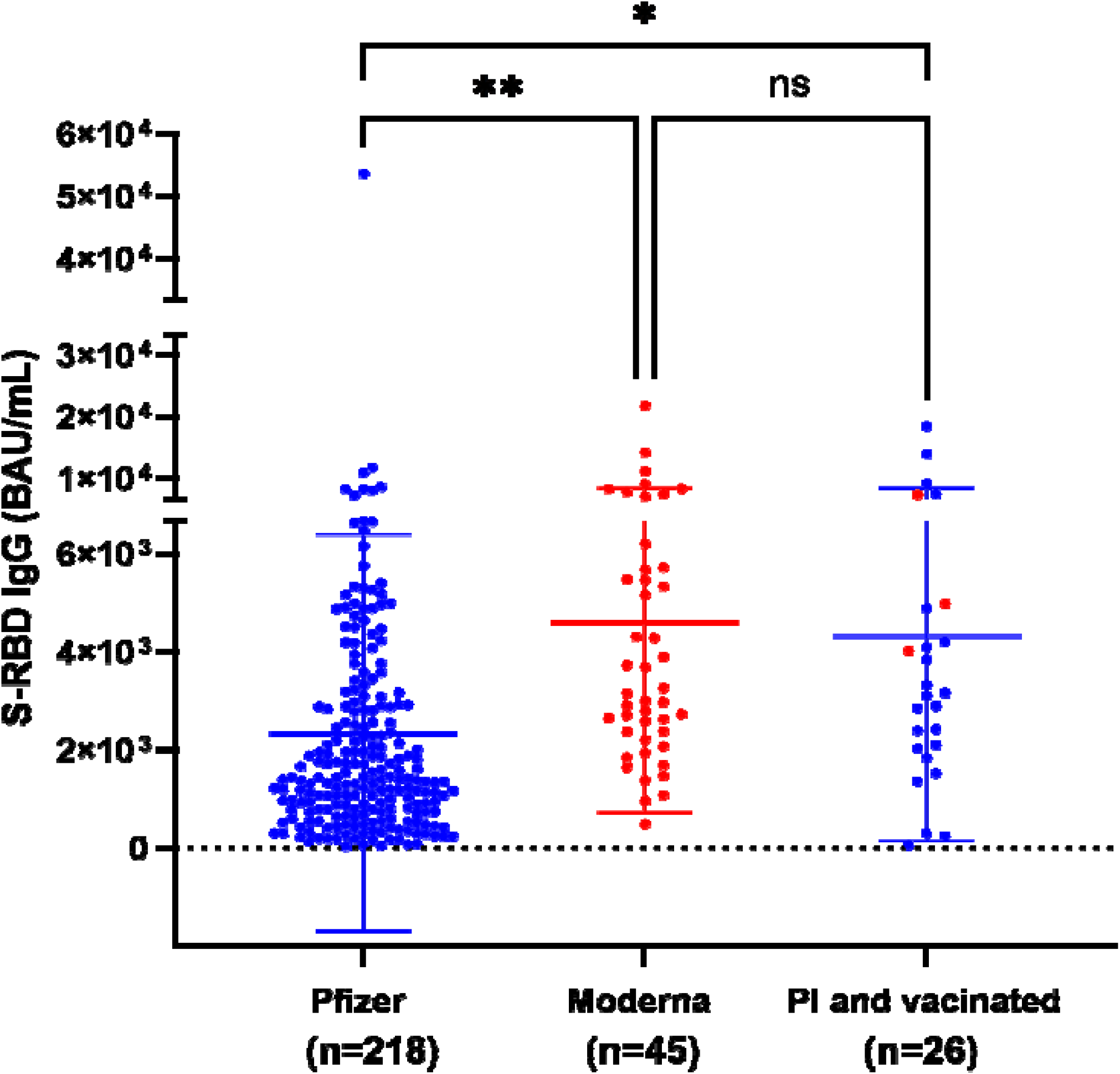
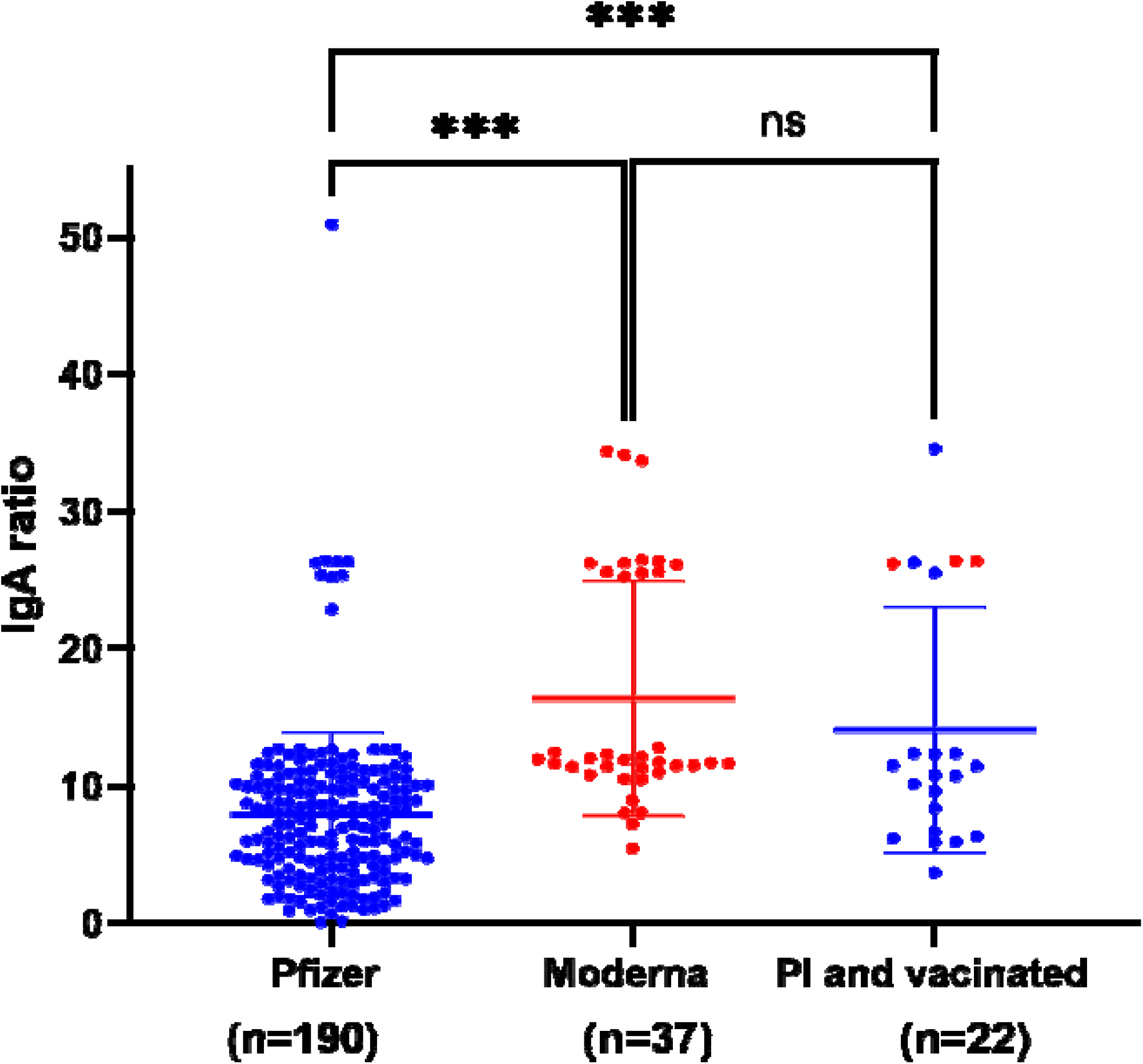
Antibody levels in mRNA vaccinated participants after 1-13 (median=6) weeks of receiving two doses, and participants with prior infection with two doses of vaccine. The tests were performed using the automated analyzer Mindray Cl-900i **(A)** S-RBD Total antibodies level (AU/mL) **(B)** S-RBD IgG antibody levels (BAU/mL) **(C)** anti-S IgA antibody levels using Euroimmun ELISA. ns p>0.05, * p≤ 0.05, ** p≤ 0.01, *** p ≤ 0.001.

#### 3.2.2 S-RBD IgG response

S-RBD IgG response was detected in all naïve-BNT162b2, naïve-mRNA-1273, and PI vaccinated groups (Figure 1B). The mean IgG levels were 2.3×10^3^ BAU/mL (95%CI: 1.8-2.8×10^3^), 4.6×10^3^ BAU/mL (95%CI: 3.4-5.7×10^3^), 4.3 ×10^3^ BAU/mL (95%CI: 2.6-5.9×10^3^), respectively. The naïve-mRNA-1273 group produced significantly higher IgG levels compared to the naïve-BNT162b2 vaccinated group (2-fold, p=0.002). Interestingly, The PI group produced a higher level of S-RBD IgG than the naïve-BNT162b2 (p=0.05) but not more than the naïve-mRNA-1273 (p=0.9) vaccinated group.

#### 3.2.3 S-RBD IgA response

IgA antibodies were detected in all naïve-mRNA-1273 and PI vaccinated groups (Figure 1C). However, in the naïve-BNT162b2 group, 96.8% (184/190) were positive to anti-S IgA, 1.6% (3/190) were negative, and 1.6% (3/190) were borderline. The mean IgA ratios were 7.9 (95%CI: 7.1-8.7) in the naïve-BNT162b2, 16.4 (95%CI: 13.5-19.2) in the naïve-mRNA-1273, and 14.1 (95%CI: 10.1-18.1) in the PI vaccinated group. No significant difference in the IgA level was detected between the PI and naïve-mRNA-1273 vaccinated group (p=0.4139). However, the naïve-BNT162b2 vaccinated group produced a significantly lower IgA ratio than was observed in naïve-mRNA-1273 and the PI vaccinated group (P<0.001). Collectively, our results indicate that the mRNA-1273 vaccine induces a significantly higher IgA antibody response in the naïve and the PI vaccinated individuals compared to the BNT162b2 vaccine.

### 3.3 Age effect on antibody response

Samples from the naïve-BNT162b2 vaccinated participants were categorized in different age groups <30, 30-50, and >50 years old; the mean total antibodies level for each group was 3.7×10^3^ AU/mL, 4.7×10^3^ AU/mL, and 6.7×10^3^ AU/mL, respectively. The mean S-RBD IgG level for each group was 2.8×10^3^ BAU/mL, 2.2×10^3^ BAU/mL, and 1.5×10^3^ BAU/mL, respectively. The mean anti-S IgA levels for each group were 9.5, 7.2, and 6.4, respectively. Although there are differences in all antibody responses between the three age groups, however, this difference was not significant (P>0.05) (Figure 2).

**Figure 2.**
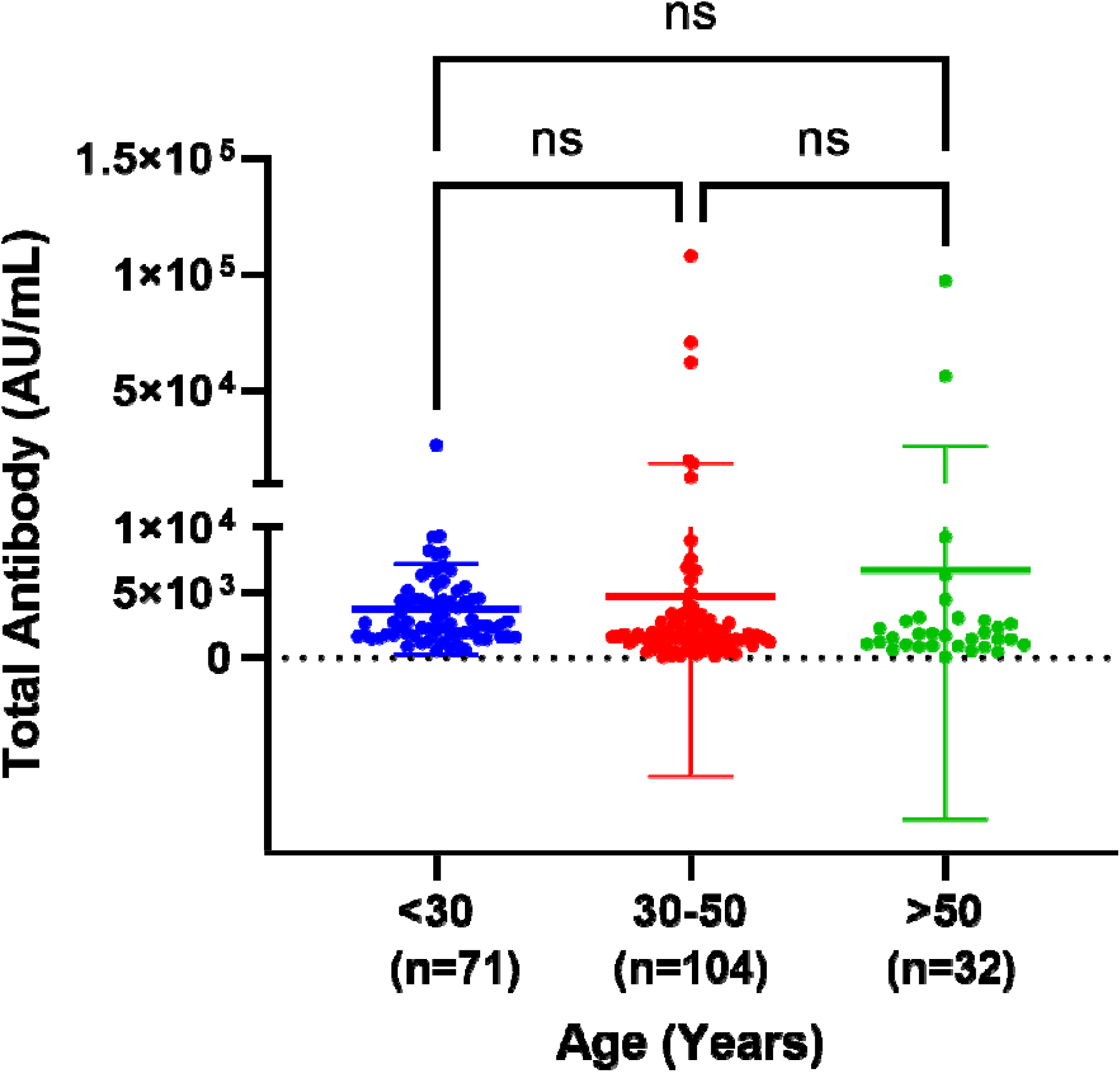
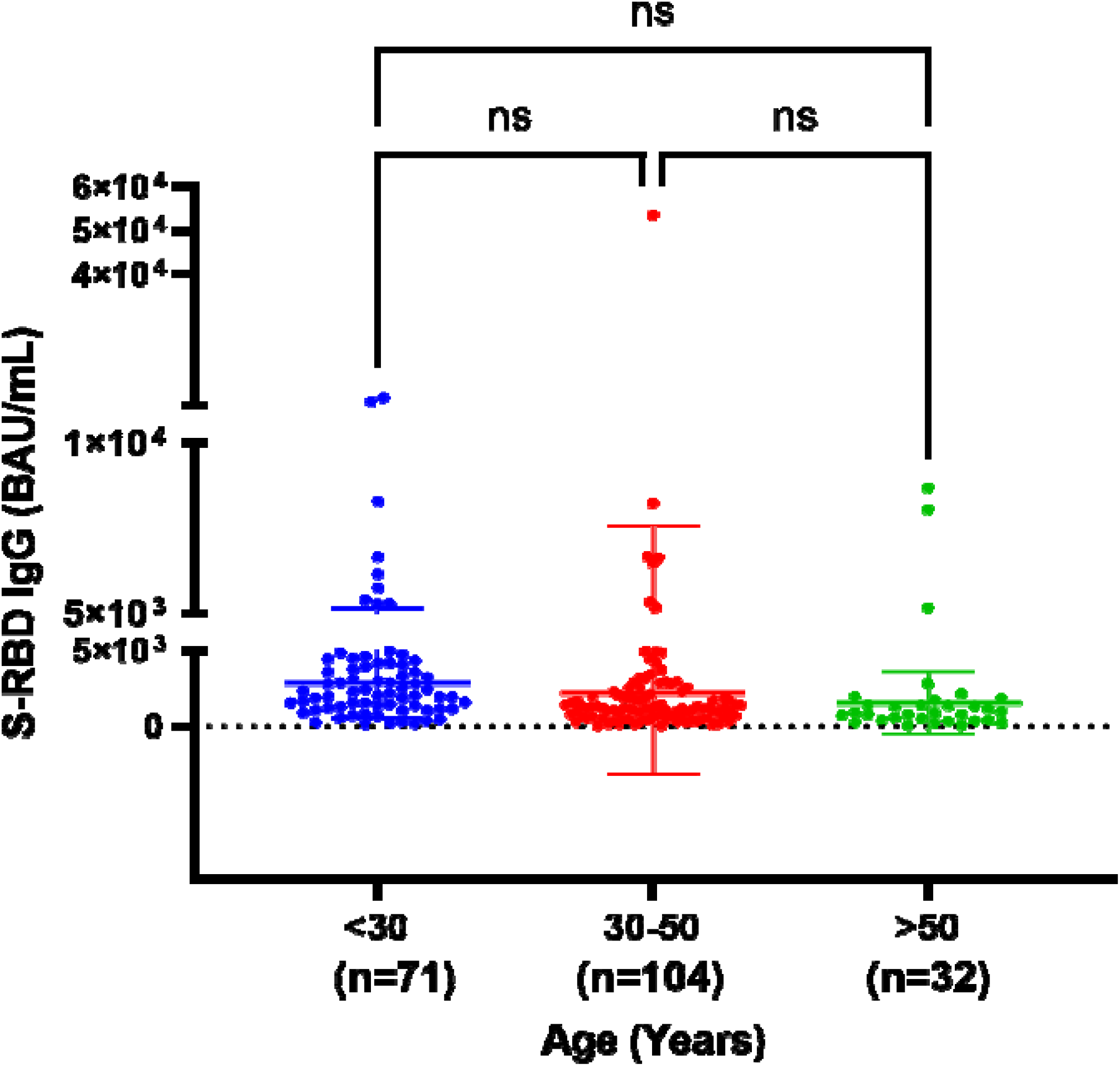
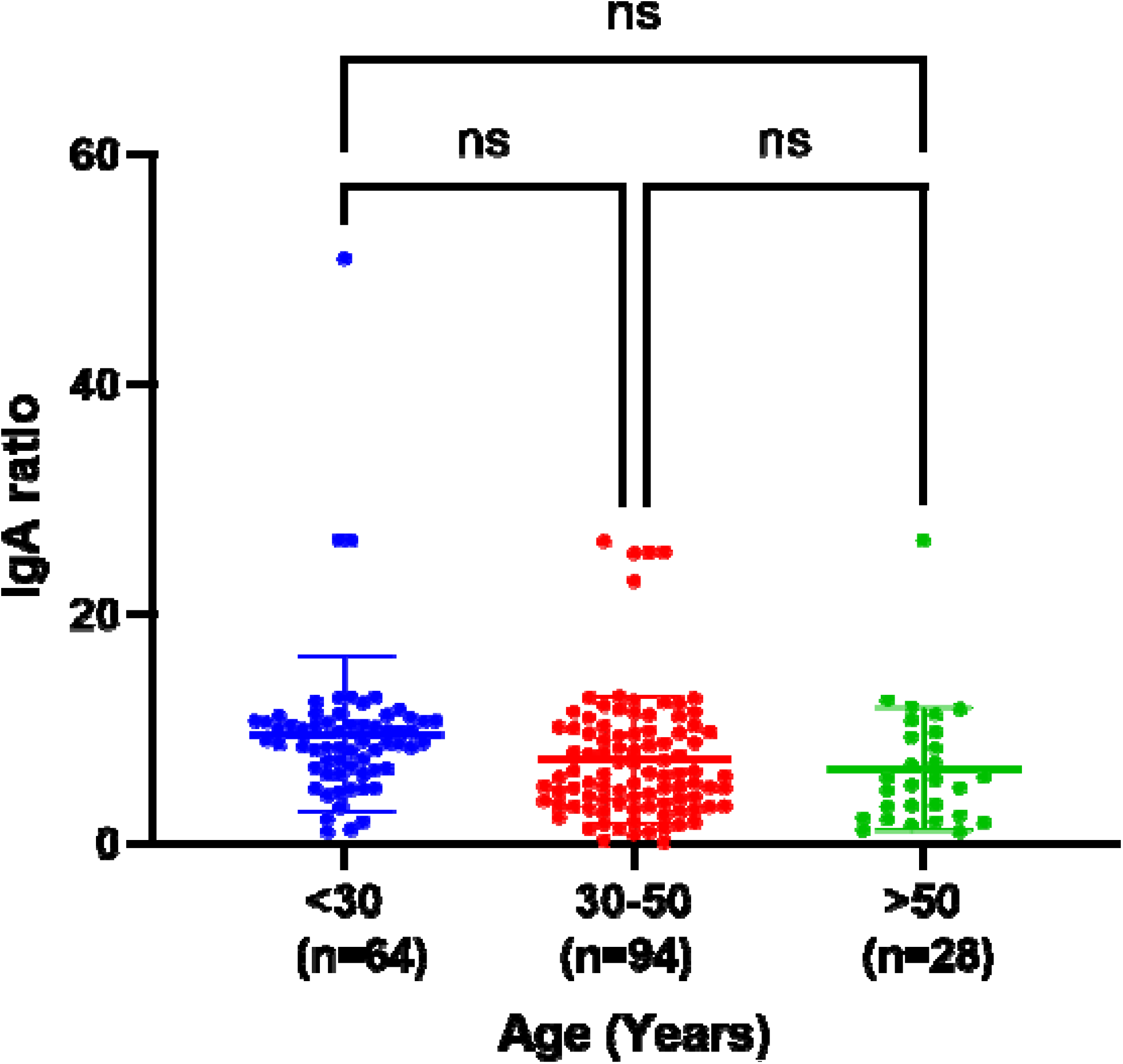
Antibody levels in BNT162b2 vaccinated participants according to age groups. (A) Total antibodies level (Au/ml) (B) S-RBD IgG antibody levels (BAU/ml). (C) anti-S IgA antibody levels. ns P>0.05.

## 4. Discussion

The mRNA vaccines BNT162b2 and mRNA-1273 offer great promise for curbing the spread of SARS-CoV-2 infection. They showed an efficacy rate of more than 94% [3, 4] and were reported to be safe. Recently we have demonstrated that the effectiveness of the mRNA-1273 shows similar levels and patterns of protection to the BNT162b2 vaccine [10]. However, the mRNA-1273 appears to be more robust against B.1.351 and provides greater protection than the BNT162b2 [5]. A relationship between neutralization level after SARS-CoV-2 vaccination and protection against COVID-19 was previously emphasized [11]. The level of the antibody response after vaccination correlates with neutralizing antibody titers, which might be clinically significant [12]. In the current study, robust antibody response was clearly observed in both mRNA vaccinated participants. All participants elicited anti-S-RBD total antibodies and anti-S-RBD IgG and almost all produced S-RBD-IgA (Figure 1). Yet, differences were observed in antibody response between BNT162b2 and mRNA-1273 groups. To our knowledge, there is only one published report with similar vaccinated group criteria that showed parallel results to ours [13]. The novelty of our study stems from testing three different parameters to measure antibody response in mRNA vaccine participants, including the IgA response, which was not previously measured [13]. Natural infection mediates viral neutralization through the production of IgA antibodies. However, little is known regarding the vaccine-induced immune response [14]. Interestingly, a recent study demonstrated that IgA dominates the early neutralizing antibody response to SARS-CoV-2 [15] and this could be clinically significant for protection. We specifically tested the total and the IgG response not to the whole spike S protein but only to the spike protein S-receptor-binding domain (S-RBD). We expect the anti-SRBD antibodies to specifically correlate with the antibody neutralizing activity as both targeted the S-RBD.

Wheeler et al. [16] reported no differences in antibody responses (anti-S1, anti-RBD, and anti-S2) between mRNA-1273 and BNT162b after receiving the first or the second dose. Here we showed that the mRNA-1273 vaccine significantly produces higher antibody response levels for S-RBD IgG, anti-S IgA, and total antibodies compared to BNT162b2 with at least two-fold in all antibody tested parameters (Figure 1). Our study results are in agreement with several other reports [13]. For instance, a recent study demonstrated that the mRNA-1273 vaccine produces a significantly higher total antibodies level to the whole S-spike protein with the BNT162b2 vaccine in both infected and uninfected groups. Besides, this was observed across different age categories (14). The differences in the level of antibody response are potentially due to each vaccine’s formulation, dose content, and the interval between the doses. It was reported that mRNA-1273 has higher mRNA content than BNT162b2 (100µg vs. 30µg, respectively). Another factor could be the time elapsed between the first and second doses [5]. For instance, the mRNA-1273 vaccine has two doses 28 days apart, while BNT162b2 is 21 days apart. Therefore, this might have affected the build-up of immunity after vaccination.

As expected, we showed that PI participants who received two doses of mRNA vaccines produced significantly higher total antibodies titers compared to the naïve vaccinated group (Figure 1A), which is in agreement with a previous study [13]. In addition, PI vaccinated produced higher antibody levels compared to the naïve BNT162b2 vaccinated group. However, no significant difference in the levels of S-RBD IgG and anti-S IgA was observed between PI vaccinated, and naïve mRNA-1273 vaccinated individuals (Figure 1B). These results emphasize that the elicited antibodies in response to mRNA vaccines include IgA and IgM, which explains the differences in the total antibodies level (Figure 1A). In addition, our PI cohort included samples with different times of infection, which might explain the variation in antibody titers and response to the vaccine. The reason why mRNA-1273 produced higher IgA levels needs further investigation, whether this could be due to dose-effect or the duration between the two doses.

On the other hand, it was expected that there is a distinct SARS-CoV-2 viral-specific antibody response and varies based on age. Although we showed that the <30 group produces higher levels than the other two groups, however, this difference was not significant (P>0.5) (Figure 2). Some studies reported that the initial response to different SARS-CoV-2 antigens is age-dependent. For instance, Jalkanen et al. reported that after receiving the first dose of BNT162b2, the levels of antibodies were significantly lower in the old age group (>50) compared to the younger age groups. However, The difference in antibody responses disappears after receiving the second dose [17]. In fact, Wheeler et al. [16] reported the minimal effect of age and gender on antibody responses after vaccination.

In conclusion, our ongoing study showed that the antibody response induced by mRNA-1327 was approximately three-fold higher than BNT162b2. In addition, higher total antibodies titer was reported in PI vaccinated compared to naïve vaccinated participants. While both mRNA SARS-CoV-2 vaccines have high efficacy and strongly protect against infection, mRNA-1327 remains the most effective over time. The study has some limitations; other immune parameters (SRBD-IgM and the neutralizing antibodies), the durability, and the kinetics of antibodies after vaccination need further investigation to provide a complete immune response profile. Further, most of the PI group received the BNT162b2 and only few received the mRNA-1327 vaccine.

## Data Availability

The data that support the findings of this study are available upon request from the authors.

## Acknowledgment

We would like to thank Ms. Sahar Aboalmaaly, Ms. Afra Al Farsi, Ms. Reeham Al-Buainain, Ms. Samar Ataelmannan, and Ms. Jiji Paul, the laboratory technologists at the Medical Commission Laboratory Section, Ministry of Public Health, Qatar, for their help in performing the Architect® immunoassay.

